# A comparative analysis of the health, financial, equity, and cost-effectiveness impacts of maxillofacial surgery in Guinea

**DOI:** 10.1101/2021.03.24.21254058

**Authors:** Mirjam Hamer, Dennis Alcorn, Ibrahima Diallo, Fatoumata B.Y Bah, Alhassane Conde, Lancinè Traoré, Etienne Millimounou, Chelsea Peacock, Chris Glasgo, Peter E. Linz, Mark Shrime, Oumar Raphiou Diallo

## Abstract

**Background:** Non-governmental organizations (NGOs) play a substantive role in the delivery of surgical services in in low- and middle-income countries (LMICs).

Assessment of their outcomes, especially as they relate to outcomes of surgery done in country, remains limited.

**Methods:** A prospective analysis of maxillofacial surgery in Guinea. Outcomes of interest were changes in patient health, subjective well-being, and financial status; hardship financing and catastrophic expenditure; equitable distribution of surgical access; and cost-effectiveness.

**Results:** We followed 569 patients requiring maxillofacial surgery in Conakry, Guinea, 114 of whom got care at local university hospitals, and 455 of whom got their care with Mercy Ships, a surgical NGO. Patients were followed for between three months (local) and one year (NGO). All patients reported significant improvement in objective and subjective measures of health and in financial status. Approximately half had to borrow and sell to get care, with NGO patients borrowing less, on average. However, NGO patients faced more risk of catastrophic expenditure (41.2% vs. 28.1%, *p* < 0.001). NGO patients were significantly poorer, whether financial status was measured by asset wealth or monthly income (p < 0.001). Finally, surgical care by the NGO was cost effective.

**Conclusions:** In a prospective analysis of surgical patients in an LMIC, we find that surgery improves health and financial well-being. NGOs may be able to reach patients who would not be able to get care through their local system; however, this comes at a cost of increased initial financial risk. Finally, NGO-based surgical care is cost-effective.

## Introduction

Surgical care is crucial to well-functioning health systems. Every year an estimated 16.9 million people die from conditions requiring adequate surgical care, representing 32.9% of all deaths worldwide.^1^ Nearly 70% of the global population cannot access surgery if they need it. In addition, of patients who get surgery every year, 81lJmillion are forced into poverty by its costs. In fact, out-of-pocket (OOP) payments for health care are the predominant form of financing in many regions. ^2,3^

Non-governmental organizations (NGOs) play a large role in the delivery of surgical care in low- and middle-income countries (LMICs). In 2016, Ng-Kamstra *et al*. (2016) cataloged 403 NGOs providing surgery in LMICs.^4^ Although these NGOs aim to fill the gap of access to safe, affordable and timely surgery for underserved populations, they have come under increasing critique. There is a wide acknowledgement that solely providing surgery is insufficient, and that simply reporting the number of surgeries done are not satisfactory to measure the success of these initiatives.^5–7^Although some reviews have shown the positive impact of surgical missions, the level of evidence for this remains low. Botman *et al*. (2021) stated that > 90% of experienced surgical mission workers from various NGOs emphasized the importance of long-term follow-up (>6 months) and the reporting of it.^8^ In addition, NGO surgery can be costly; the cost-effectiveness of such missions must be examined.^9,10^

As a result, there have been calls for NGOs to take responsibility for measuring and reporting their outcomes and impact.^11^ In this paper, we propose an evaluation of the impact of surgical care that goes beyond measuring volume. Instead, we propose a multiattribute framework, evaluating the impacts of care on the health and financial well-being of patients, the equitable distribution of surgery within a region, and the cost-effectiveness of two models of care delivery. This holistic metric will be applied to patients presenting with maxillofacial, head and neck surgical disease in Guinea, West Africa.

## Methods

### Geographic and delivery context

Guinea is a West African country of 12.4 million people within 95,000 square miles. Fourteen percent of the country lives in the capital, Conakry. There are 8.3 physicians and 12.4 nurses per 100,000 people in the population.^20^

Mercy Ships is a charitable NGO providing specialized, elective surgery using hospital ships. The hospital aboard the flagship, the m/v *Africa Mercy*, has been described elsewhere in detail.^12–14^ The ship was docked in Conakry for a ten-month field service that began in August 2018. Although surgeons come to the ship from seven specialties, only the maxillofacial / head and neck service spans the entire field service. This study follows only these patients.

Two University Teaching Hospitals in Conakry—Donka and Ignace Deen—provide local maxillofacial and head and neck care.^15^ One of the authors (ORD) is the single fellowship-trained maxillofacial surgeon in the country. He works at both hospitals, with a team of 26 dentists and surgical trainees. Patients seen at Donka and Ignace Deen served as the comparator group.

### Disease context

Oral disease is estimated to affect more than 4 billion people annually, disproportionately among people of lower socioeconomic status.^16^ Although treatment of oral conditions often lies within the purview of a dentist, delays in access to care can lead to delayed presentation and subsequent reliance on oral and maxillofacial (OMF) surgeons. Among conditions addressed by OMF surgeons are complications of dental disease, infections, benign maxillofacial cysts, tumors and malformations, oral cavity cancer, orofacial clefts, and OMF trauma, among other conditions.^17^

OMF conditions need not be malignant to exert a toll. Although data on prevalence and burden of OMF conditions in LMICs are scarce, it is likely that they play an important part in the global burden of surgical disease. The effects of OMF conditions can be devastating for patients and can impact the position they have in society. In addition to the functional limitation these conditions cause, social isolation is often a common consequence. These factors can then directly or indirectly lead to downstream financial hardship.^17^

### Definition of Impact

Multiple evaluative frameworks have been proposed in global health.^18,19^ In this study, we have chosen to use a holistic, extended cost-effectiveness analysis framework (**Table 1**), modeled after the WHO’s Universal Health Coverage.^20^ This framework has been used in multiple previous modeling studies in global surgery.^21,22^

**Table 1:**
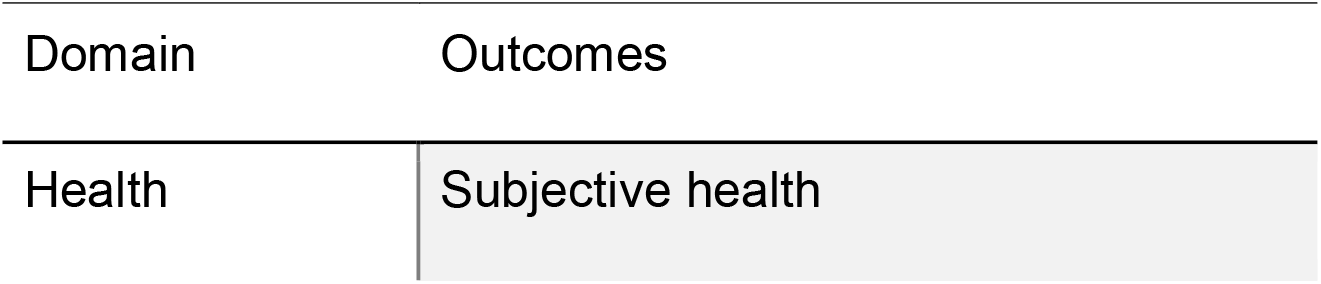
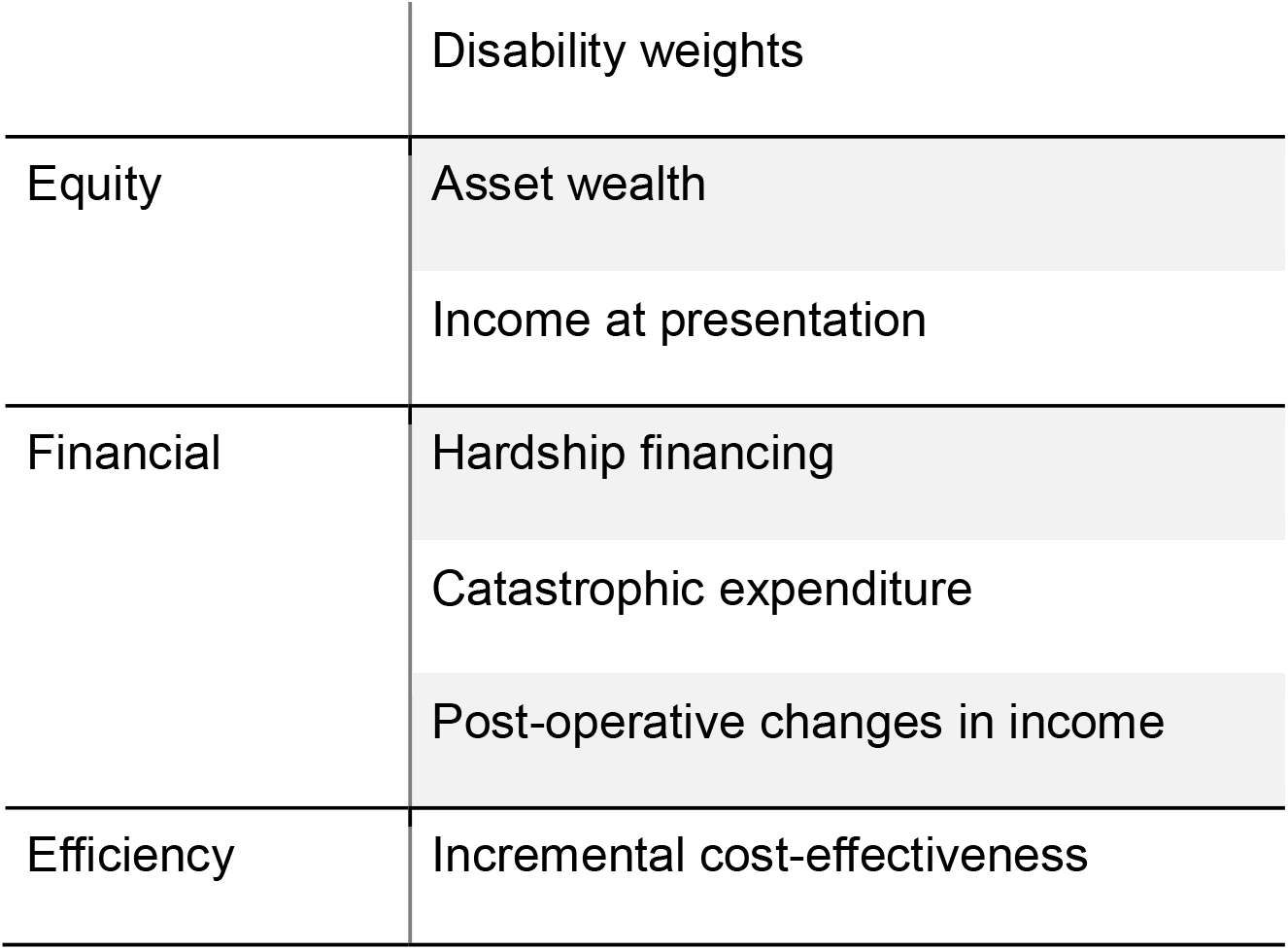
Domains of impact

| Domain | Outcomes |
| --- | --- |
| Health | Subjective health |
|  | Disability weights |
| Equity | Asset wealth |
|  | Income at presentation |
| Financial | Hardship financing |
|  | Catastrophic expenditure |
|  | Post-operative changes in income |
| Efficiency | Incremental cost-effectiveness |

All patients were surveyed on admission to the hospital and, by telephone, at three months post-operatively. Patients presenting to Mercy Ships were also interviewed by telephone at twelve months post-operatively. Survey responses were entered into REDCap Cloud, a cloud-based graphical user interface data capture and management software. Surveys are available in the **Appendix**.

### Patient selection

All patients presenting with maxillofacial and head and neck conditions to Mercy Ships and to three local institutions (Donka University Teaching Hospital, Ignace Deen University Teaching Hospital, and the private clinic of ORD) were approached for recruitment. Consent was obtained in the patient’s local language. If patients were children, their parents were asked to participate.

### Health

All diagnoses were assigned an International Classification of Diseases-10 (ICD-10) code, with the maximum granularity their written diagnoses allowed. For the purposes of analysis, these conditions were then grouped into their top-level ICD-10 codes.

Patients were asked a single subjective question: “On a scale from 1 to 10, where 1 is worst and 10 is best, how healthy do you feel that you are?”

Patients then answered the 12-question WHO Disability Assessment Schedule 2.0 (WHO-DAS).^23^ Their responses were converted to a disability weight using the methods recommended by Salomon, *et al* (2003).^24^ The disability weight ranges from 0, representing no disability (or perfect health) to 1, representing maximum disability (or death).

The WHO-DAS questions cover six domains of functioning: cognition, mobility, self-care, interaction with others, activities of daily living, and community engagement. Sub-analyses were performed to assess changes across these individual domains.

Disability-adjusted life years were calculated based on the formula proposed by Murray, et al (1997)^25^:

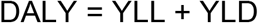

For all ICD-10 codes, a systematic literature review was conducted to determine whether any premature mortality was associated with that condition. The literature review identified any paper demonstrating increased odds of mortality or decreased survival with the identified conditions. If multiple papers reported increased mortality, the lowest estimate was taken to maintain *a fortiori* assumptions.

The World Health Organization’s 2016 life table for Guinea^26^ was used to calculate predicted life expectancy for each patient based on their age (LE_ideal_), as well as predicted life expectancy adjusted for their condition’s increased risk of death, if applicable (LE_actual_).

Disability weights, calculated from pre- and post-operative WHO-DAS answers, as above, were combined with these life expectancies to calculate total averted DALYs per patient as follows:

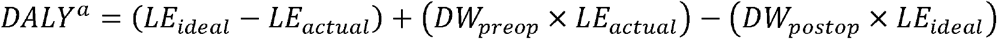

### Equity

We used a previously validated method for converting asset wealth into wealth quintiles to categorize patients into five quintiles—the poorest 20% to the richest 20%.^27^ The asset wealth survey has been previously published.^27^ Quintile distribution was compared for local vs. Mercy Ships patients and for Mercy Ships patients vs. standardized urban wealth quintiles for Guinea as a whole.

### Financial outcomes

Patients were also asked about their income over the last month. This was converted to US dollars using the prevailing exchange rate.^28^ Hardship financing and financial catastrophe were measured. For the former, respondents were asked if they had to borrow money or to sell assets to pay for care. If they did borrow money, the amount they borrowed was obtained.

Catastrophic expenditure was defined as any expenditure related to their surgical care that was more than 10% of their overall yearly household expenditure.^29^ All surveys are available in the **supplementary Appendix**.

### Cost effectiveness

This paper does not assume that, without the presence of Mercy Ships, patients would not be getting surgical care. As a result, our primary cost-effectiveness outcome is the incremental cost-effectiveness ratio (or ICER), calculated as:

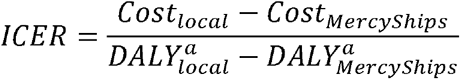

For comparison with other, previously published NGO papers, an average cost-effectiveness ratio is also reported:

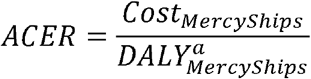

Costing data was provided from Mercy Ships using a full-accounting, micro costing approach. This costing included all fixed and variable costs accounted to the maxillofacial service on Mercy Ships, including the calculated value of lost wages for volunteers. Because fixed costs were not available for the local hospitals, they were estimated to be the cost of equipping and staffing a single operating room based on previously published data.^30^ As this was the largest source of uncertainty in the cost-effectiveness calculation, it was subjected to sensitivity analyses.

### Statistical analysis, ethics, and funding

For any non-normally distributed data, Kruskall-Wallis tests were used to compare groups. For any normally distributed data, Student’s *t*-tests were used. Analyses were performed in R v3.6.3. This study was approved by the Partners Health System IRB and by the Mercy Ships Research Ethics Council. Funding for this study was supplied by Mercy Ships.

## Results

### Patient Demographics and follow-up

Patient demographics can be found in **Table 2**

**Table 2:** Demographics of surgical patients in Guinea

|  | Local patients | Mercy Ships patients | *p* |
| --- | --- | --- | --- |
| N | 114 | 455 |  |
| Age, mean (SD) | 29.01 (14.68) | 29.49 (18.92) | 0.802 |
| Male, N (%) | 76 (66.7) | 212 (46.6) | <0.001 |
| Married, N (%) | 38 (33.3) | 193 (43.9) | 0.054 |
| Number of children, mean (SD) | 0.52 (1.26) | 2.16 (2.81) | <0.001 |
| Number of years in school, mean (SD) | 7.10 (6.51) | 4.26 (5.33) | <0.001 |
| Inpatient LOS, mean (SD) | 7.76 (10.14) | 6.47 (5.75) | 0.072 |

Compared with Mercy Ships patients, a significantly higher proportion of local patients were male (66.7% vs 46.6%, *p* < 0.001). On average, Mercy Ships patients had more children (2.16 vs. 0.52, *p* < 0.001) and fewer years of schooling (4.26 vs. 7.10, *p* < 0.001).

ICD-10 code breakdowns for both groups of patients can be found in the **supplementary Appendix**.

At three months, 86% of Mercy Ships patients completed at least part of the follow-up survey, while 95% of local patients did. At 12 months, 89% of the original Mercy Ships cohort completed at least part of the overall survey.

### Health

Patients’ subjective health significantly improved in both cohorts, as can be seen in **Figure 1**. Differences from preoperative responses are significant in both groups (*p* < 0.001).

**Figure 1:**
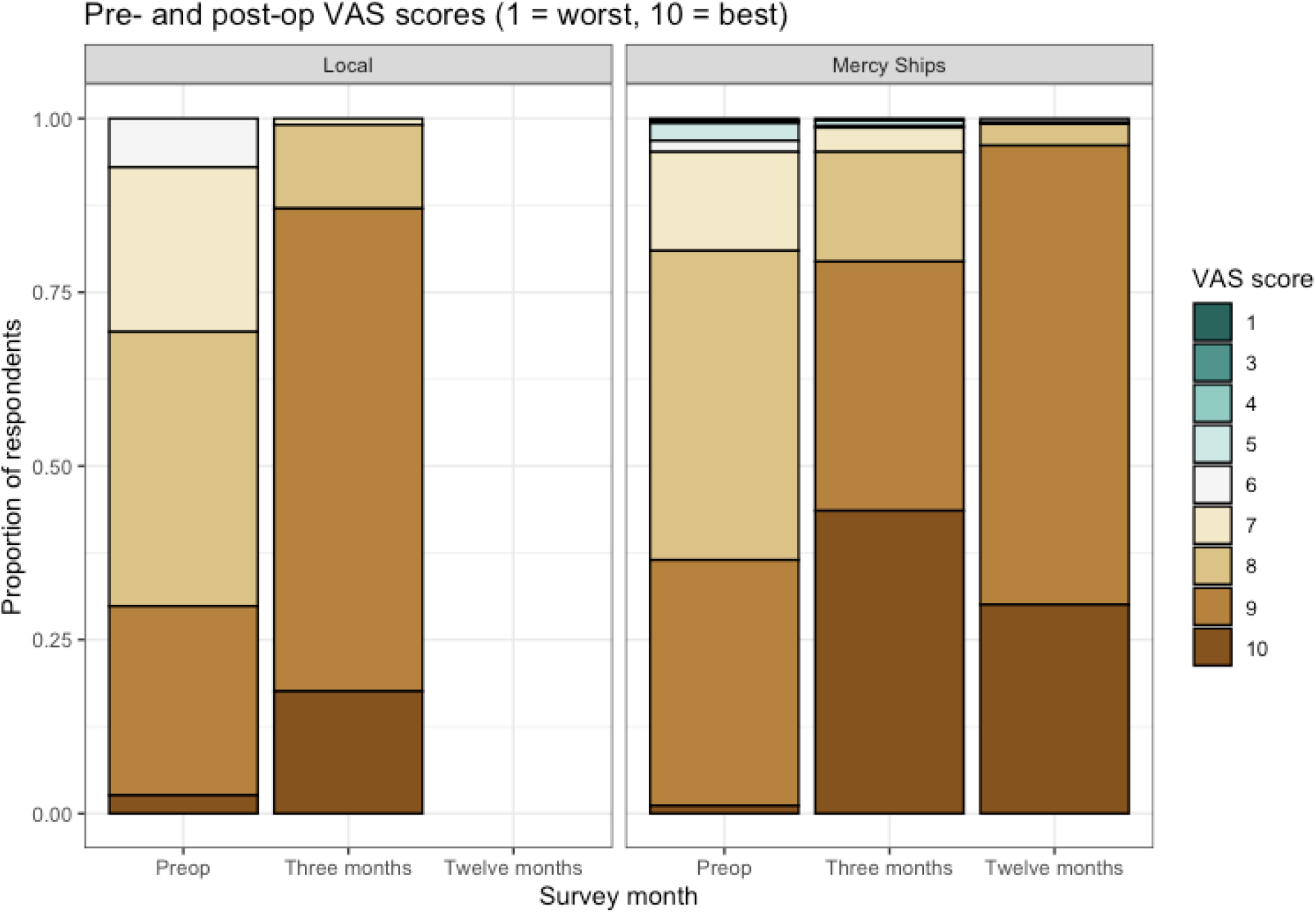
Subjective health. VAS = visual analog scale. 1 represents the worst health and 10 represents the best.

Calculated disability weights among respondents also significantly improved post-operatively (*p* < 0.001 for Mercy Ships patients, *p* = 0.001 for local patients, **Figure 2**). The improvement was sustained over the first post-op year in Mercy Ships patients. The difference in calculated disability weight among the two groups was not statistically significant (*p* = 0.172).

**Figure 2:**
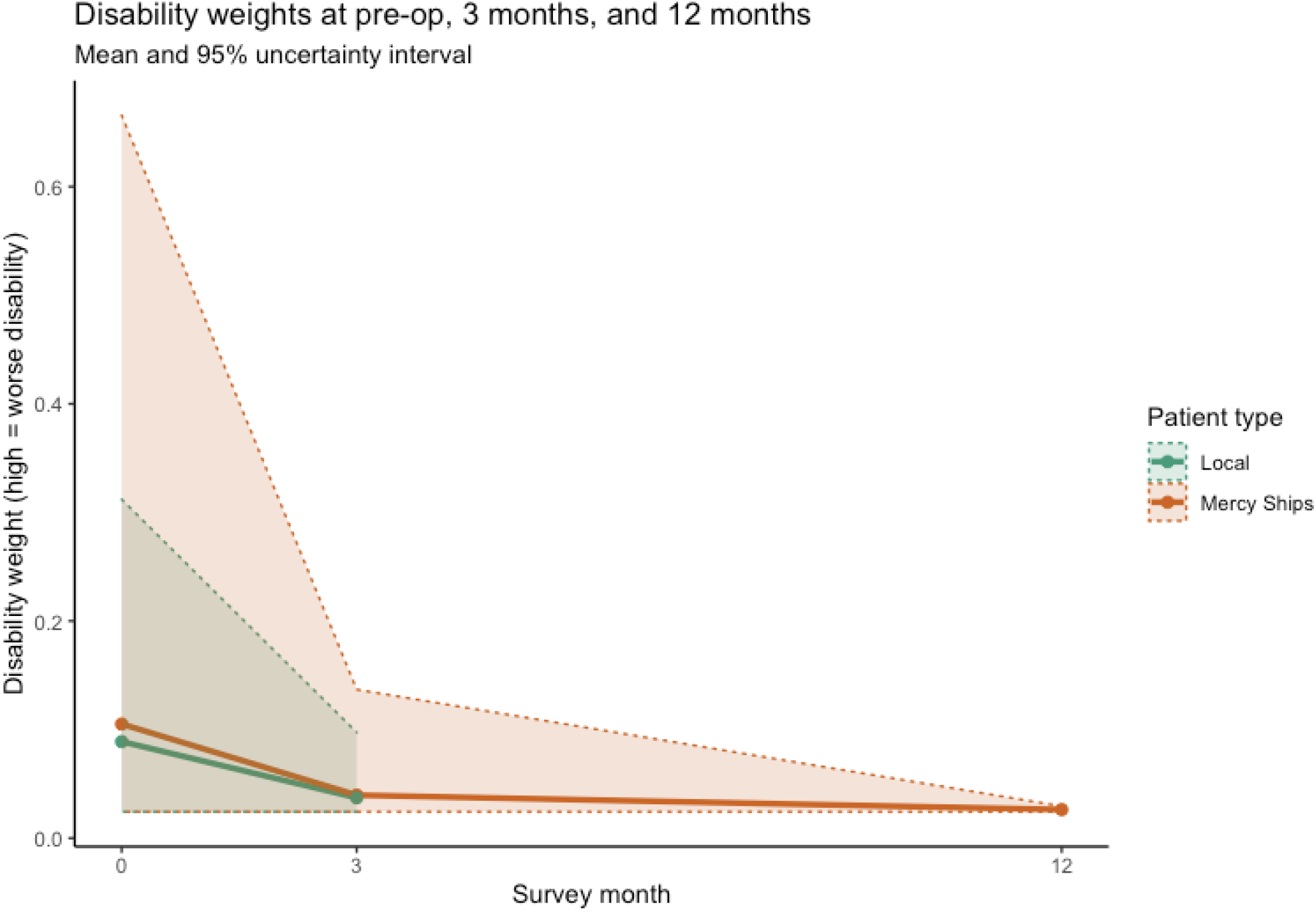
Disability weights. 0 = perfect health. 1 = death.

Of the six domains within the WHO-DAS survey, Mercy Ships patients reported the most improvement along the domain of community engagement.

### Equity and financial status

Mercy Ships patients have significantly less asset wealth than surgical patients presenting to local hospitals (*p* < 0.001, **Figure 3**). In addition, their starting monthly incomes were significantly lower (USD 221 vs. 101, *p* < 0.001). Postoperatively, both groups’ incomes improved (**Figure 4**). The improvement was significant in both local patients (*p* = 0.015) and significant among Mercy Ships patients (*p* < 0.001).

**Figure 3:**
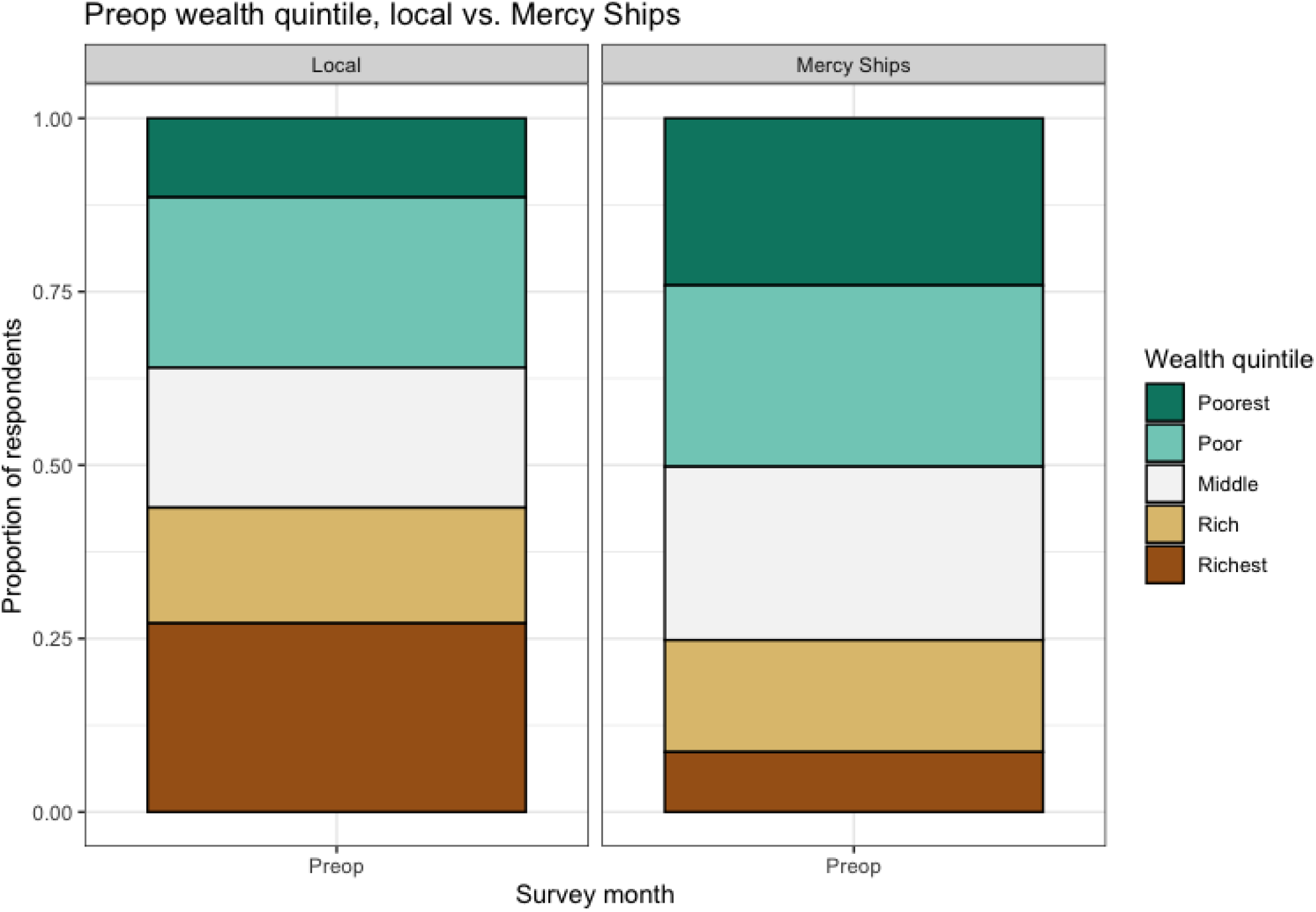
Proportion of respondents whose asset wealth places them into each wealth quintile in Guinea

**Figure 4:**
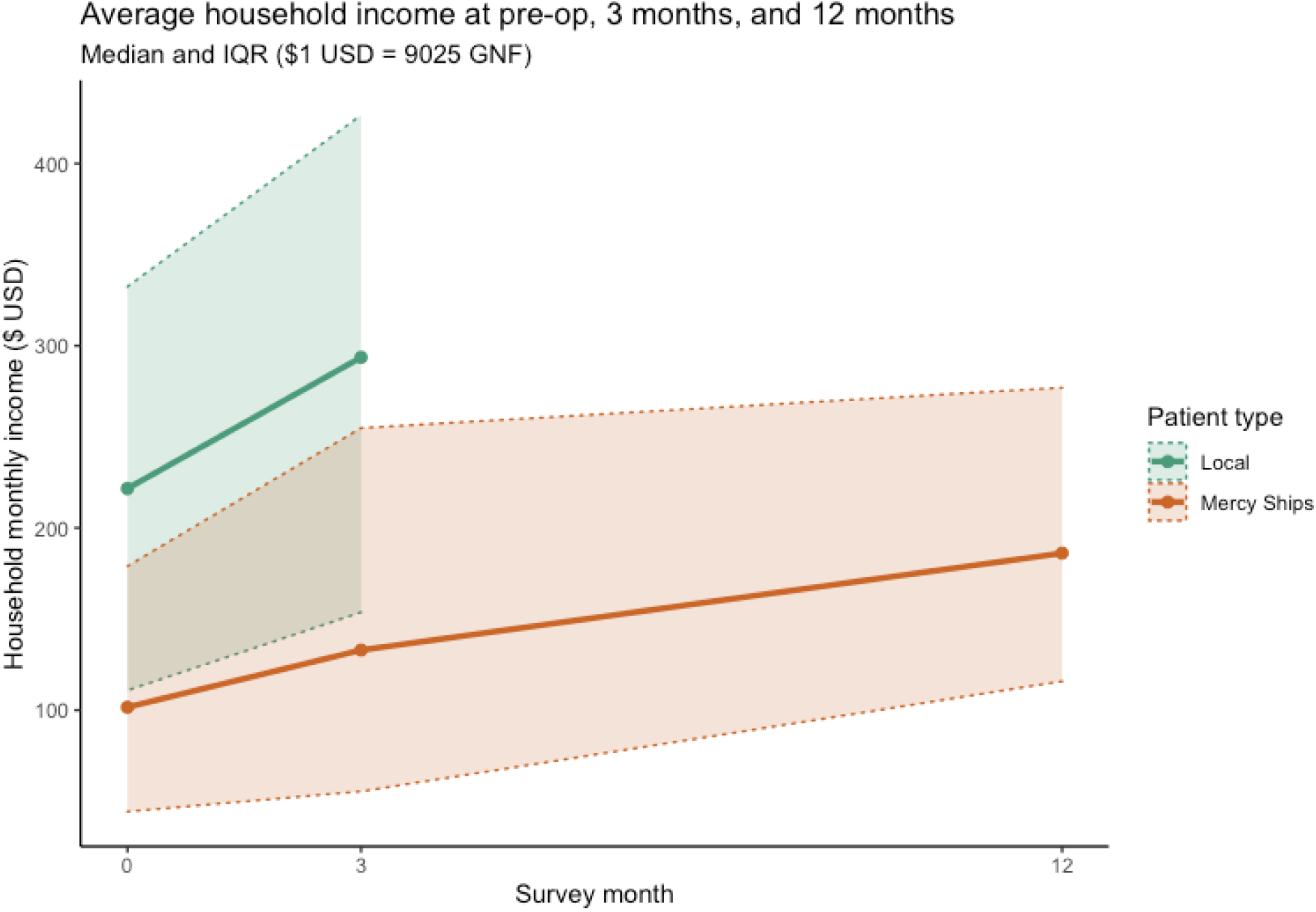
Income, in USD

### Financial risk

Both groups of patients had to borrow money or sell belongings to access surgical care at similar rates (48.3% for Mercy Ships patients, 46.2% for local patients, *p* = 0.841). However, among patients who had to borrow money, Mercy Ships patients borrowed significantly less (USD 54 vs 280, *p* < 0.001).

Both groups of patients faced catastrophic expense, with a higher likelihood of catastrophic expense among Mercy Ships patients (41.2% vs. 28.1%, *p* < 0.001).

### Cost-effectiveness

Per-patient mean total costs and DALYs averted for maxillofacial surgery at local hospitals and on Mercy Ships can be found in **Table 3**.

**Table 3:** Cost-effectiveness analysis. ICER = incremental cost-effectiveness ratio.

|  | Cost (USD) | DALYs averted | ICER |
| --- | --- | --- | --- |
| Local | 7109.08 | 2.8286 | (Weakly Dominated) |
| Mercy Ships | 11,175.42 | 4.9856 | 1885.20 |

Average cost-effectiveness ratios show a cost per DALY averted by Mercy Ships of USD 2,242 and by local hospitals of USD 2,513. Surgery on Mercy Ships weakly dominates surgery performed at local institutions. However, this dominance is sensitive to the total per-patient cost of surgery at local institutions. When the total cost, inclusive of fixed costs, at local hospitals drops below USD 5141.64, surgery at local hospitals is no longer dominated.

## Discussion

In this paper, we show that a multifactorial impact evaluation with good long-term follow-up is not only feasible for NGOs to perform, but also that doing so highlights the various effects these organizations can have on surgical patients. Additionally, we compared the effect of an NGO with the counterfactual—what exists in the surgical system outside the work of the NGO. We find that treating surgical disease increases health, improves disability, and increases patients’ abilities to return to the workforce. We also find that the examined NGO was able to access poorer patients than routinely present for care at local institutions. Both local and NGO patients face financial hardship: all patients borrow or sell belongings to get care at the same rate, while NGO patients also face nearly double the rate of catastrophic expense. Finally, we find that surgery by the NGO, despite the funds needed to run it, is cost-effective.

Long-term follow-up has been difficult in global surgery. For example, Bermudez *et al*. (2010) attempted to follow up 4086 patients who had surgery for cleft lip and palate by Operation Smile. Only 20% (812 patients) returned for a 6-9-month postoperative evaluation.^31^ Other studies report similar follow up rates, ranging from 17-36%.^31–33^ We believe the increased follow-up in this study (86-93%) is attributable to strong relationships between the NGO and the local health system, and a dedicated team of local employees that were part of the study. For patients presenting to local hospitals, medical doctors in the hospitals have been key to follow-up. Finally, follow-up over the phone proved sufficient for the data collection done here. Phone follow-up likely leads to decreased attrition because patients do not have to return to the hospital.

### Health

Tracking health requires more than simply documenting clinical outcomes. Numerous papers talk about the impact disfigurement can have on these patients and how they are socially ostracized.^34,35^ (Self-) exclusion from the community, and therefore a loss of their social network is not uncommon. We show that, perhaps unsurprisingly, objective and subjective health are improved post-surgically, irrespective of the institution doing the surgery. We also show that this improvement is sustained for up to a year post-operatively.

Of particular interest is the fact that the largest improvement in the WHO-DAS 2.0 domains for maxillofacial patients was seen in the domain of community engagement. This suggests that surgery impacts patients on more than just a physical level, and that relieving the burden of head and neck disease has positive sociological externalities.

### Equity and Financial Risk

Lack of access to surgery falls heaviest on the poor. We show that surgery offered by the NGO reaches poorer people than would otherwise be able to access care at local hospitals, irrespective of how poverty is measured (i.e., asset wealth vs. income). There are knock-on effects, once again, of surgical access: post-operatively, patients seen in either setting show a statistically significant increase in their income. Taken together, these two findings suggest that charitable surgery may reach poorer patients and offer them improved financial outcomes.

Patients continue to face financial hardship, however. Almost half of the patients in either group had to borrow money to get care, even though the NGO offered surgery for free. However, the amount of money borrowed was significantly less for patients that obtained surgical care with the NGO. Catastrophic expenditure—defined as an expense of more than 10% of an individual’s income—is seen more often with NGO patients than with local patients. Although it is likely that this is because the NGO attracts poorer patients, a more thorough exploration is needed.

Nonetheless, other studies have shown a much higher incidence of catastrophic expenditures for surgery. For example, in Morocco, 88% of women in the poorest quintile who need a C-section will experience financial catastrophe.^36^ Similarly, Keya *et al*. (2018) state that for women with obstetric fistula, opportunity cost presents as an insurmountable barrier to accessing fistula repair.^37^

Finally, the gender distribution among patients presenting for care at the NGO rather than at local hospitals is different (46.6% male NGO patients vs. 66.7% male local system patients). Women tend to have higher barriers to surgical care than men do in sub-Saharan Africa, including things like a lack of access to suitable transport or to financial resources.^37–41^

### Cost-effectiveness

Cost-effectiveness in global surgery can be contentious. This is not the first paper to present a cost-effectiveness analysis of surgery.^42,43^ Many of these analyses report the total cost of NGO surgery, divide it by the total DALYs averted, to present an average cost-effectiveness ratio. This sort of analysis is predicated on a problematic assumption: that if the NGO were not there, surgery would never have happened.^44^

Instead, the recommended way to perform these analyses is to calculate the marginal cost and marginal effectiveness of the NGO—in other words, how much added benefit does the NGO provide over care at local hospitals and is that added cost worth it. This is called an “incremental cost-effectiveness ratio” and is what we report here. We find that this ratio falls below the broadly accepted threshold of 3x GDP per capita, making this NGO a cost-effective way to deliver surgery in Guinea. However, generalizability to other NGOs must be done with caution.

### Limitations

As with all studies, this has limitations. First, it was performed by one NGO in one country. Therefore, generalization should be done with caution.

Second, DALY calculations required an estimate of premature mortality, which was obtained through a review of the literature. Most retrieved papers were written in the context of a high-income country, resulting in a likely under-representation of premature mortality. This weakness would bias our analysis away from cost-effectiveness, however, giving some comfort to the fact that the reported incremental cost-effectiveness ratio is an upper limit.

Finally, the questionnaires were conducted by employees from the NGO, therefore the answers could be prone to bias. While a thorough explanation was provided, with special focus on assuring the patient that their answers would not affect their subsequent care, responses might have been given with mind toward social desirability.

Despite these limitations, this study has strengths. It is unique in its approach toward impact evaluation. It defines impact broadly, focusing not just on health but on wealth, equity, and value. In addition, it compares this to the impact surgery has in a local context. It allows for evaluation of an NGOs program and gives the opportunity to improve programmatic activities for the benefit of the patients.

The survey tools are available for use by other NGOs. It is our hope that this paper spurs the global surgery community to take responsibility for our outcomes—positive and negative—to commit to long-term follow-up, and finally to be transparent in the effects that their interventions have on patients.

## Data Availability

Data are not publicly available

## References

1. Lee JS, Roser SM, Aziz SR. Oral and Maxillofacial Surgery in Low-Income and Middle-Income Countries. Oral and Maxillofacial Surgery Clinics of North America 2020;32(3):355–65.

2. Shrime MG, Dare AJ, Alkire BC, O’Neill K, Meara JG. Catastrophic expenditure to pay for surgery worldwide: A modelling study. The Lancet Global Health 2015;3(S2):S38–44.

3. Shrime MG, Dare A, Alkire BC, Meara JG. A global country-level comparison of the financial burden of surgery. British Journal of Surgery 2016;103(11):1453–61.

4. Joshua S. Ng-Kamstra, Johanna N. Riesel, Sumedha Arya, et al. Surgical Non-governmental Organizations: Global Surgery’s Unknown Nonprofit Sector. World Journal of Surgery 2016;40:1823–41.

5. Hendriks TCC, Botman M, Rahmee CNS, et al. Impact of short-term reconstructive surgical missions: A systematic review. BMJ Global Health. 2019;4(2):1176.

6. Sykes KJ. Short-term medical service trips: A systematic review of the evidence. American Journal of Public Health. 2014;104(7).

7. Wagner K, Schwaitzberg SD, Shah Z, Joshi G, Kang J. Designing a set of safety standards for surgical short term medical missions. Global Surgery 2017;3(2):1–6.

8. Botman M, Hendriks TCC, Keetelaar AJ, et al. From short-term surgical missions towards sustainable partnerships. A survey among members of foreign teams. International Journal of Surgery Open 2021;28:63–9.

9. Bido J, Ghazinouri R, Collins JE, et al. A Conceptual Model for the Evaluation of Surgical Missions. The Journal of bone and joint surgery American volume 2018;100(6):e35.

10. Shrime MG, Sleemi A, Ravilla TD. Charitable platforms in global surgery:A systematic review of their effectiveness, cost-effectiveness, sustainability, and role training. World Journal of Surgery. 2015;39(1):10–20.

11. Zitzman E, Berkley H, Jindal RM. Accountability in global surgery missions. BMJ Global Health. 2018;3(6):1025.

12. Lin BM, White M, Glover A, et al. Barriers to Surgical Care and Health Outcomes: A Prospective Study on the Relation Between Wealth, Sex, and Postoperative Complications in the Republic of Congo. World Journal of Surgery 2016;1–10.

13. Shrime M, Hamer M, Mukhpadhyay S, et al. Effect of removing the barrier of transportation costs on surgical utilisation in Guinea, Madagascar and the Republic of Congo. BMJ Glob Health 2017;2.

14. White MC, Hamer M, Biddell J, et al. Facilitating access to surgical care through a decentralised case-finding strategy: experience in Madagascar. BMJ Global Health 2017;2(Suppl 4):e000427.

15. Chughtai M, Loua TO, Beavogui K, Qureshi AI. Neurological surgery in Guinea, west Africa. Journal of vascular and interventional neurology 2015;8(1.5):S12–6.

16. James SL, Abate D, Hassen Abate K, et al. Global, regional, and national incidence, prevalence, and years lived with disability for 354 diseases and injuries for 195 countries and territories, 1990â€”2017:a systematic analysis for the Global Burden of Disease Study 2017. 2018.

17. Reddy CL, Patterson RH, Wasserman I, Meara JG, Afshar S. Oral and Maxillofacial Surgery: An Opportunity to Improve Surgical Care and Advance Sustainable Development Globally. Oral and Maxillofacial Surgery Clinics of North America. 2020;32(3):339–54.

18. Shekelle PG, Maglione MA, Luoto J, Johnsen B, Perry TR. Global Health Evidence Evaluation Framework [Internet]. Rockville (MD): Agency for Healthcare Research and Quality (US); 2013 [cited 2021 Mar 22]. Available from: http://www.ncbi.nlm.nih.gov/books/NBK121291/

19. Bozorgmehr K, Saint VA, Tinnemann P. The “global health” education framework: a conceptual guide for monitoring, evaluation and practice. Globalization and Health 2011;7(1):8.

20. WHO. Universal health coverage (UHC) [Internet]. 2019 [cited 2021 Feb 15];Available from: https://www.who.int/news-room/fact-sheets/detail/universal-health-coverage-(uhc)

21. Shrime MG, Sekidde S, Linden A, Cohen JL, Weinstein MC, Salomon JA. Sustainable Development in Surgery: The Health, Poverty, and Equity Impacts of Charitable Surgery in Uganda. PLOS ONE 2016;

22. Shrime M, Verguet S, Johansson KA, Desalegn D, Jamison DT, Kruk ME. Task-Sharing or Public Finance for Expanding Surgical Access in Rural Ethiopia: An Extended Cost-Effectiveness Analysis. The International Bank for Reconstruction and Development / The World Bank; 2015.

23. World Health Organization. WHO Disability Assessment Schedule (WHO-DAS 2.0) [Internet]. 2015 [cited 2021 Feb 15];2–153. Available from: https://www.who.int/standards/classifications/international-classification-of-functioning-disability-and-health/who-disability-assessment-schedule

24. Salomon JA, Mathers C, Chatterji S, Sudana R, Ustun TB, Murray CJL. Health systems performance assessment. 2003.

25. Murray CJL, Acharya AK. Understanding DALYs. In: Journal of Health Economics. North-Holland; 1997. p. 703–30.

26. World Health Organization. GHO | By category | Life tables by country - Guinea [Internet]. WHO. [cited 2021 Mar 22];Available from: https://apps.who.int/gho/data/view.main.60670?lang=en

27. Lin BM, White M, Glover A, et al. Barriers to Surgical Care and Health Outcomes: A Prospective Study on the Relation Between Wealth, Sex, and Postoperative Complications in the Republic of Congo. World Journal of Surgery 2017;41:14–23.

28. XE. XE Exchange Rates [Internet]. [cited 2021 Feb 15];Available from: https://www.xe.com/

29. O’Donnell O, van Doorslaer E, Wagstaff A, Lindelow M. Analyzing Health Equity Using Household Survey Data. The World Bank; 2007.

30. Shrime MG, Sekidde S, Linden A, Cohen JL, Weinstein MC, Salomon JA. Sustainable Development in Surgery: The Health, Poverty, and Equity Impacts of Charitable Surgery in Uganda. PLOS ONE 2016;11(12):e0168867.

31. Bermudez L, Carter V, Magee W, Sherman R, Ayala R. Surgical outcomes auditing systems in humanitarian organizations. World Journal of Surgery 2010;34(3):403–10.

32. Padmanaban V, Johnston PF, Gyakobo M, Benneh A, Esinam A, Sifri ZC. Long-Term Follow-Up of Humanitarian Surgeries: Outcomes and Patient Satisfaction in Rural Ghana. Journal of Surgical Research 2020;246:106–12.

33. Gil J, Rodríguez JM, Hernández Q, et al. Do hernia operations in African international cooperation programmes provide good quality? World Journal of Surgery 2012;36(12):2795–801.

34. Camille A, Evelyne AK, Martial AE, Denise K, Marie-Josée TA, Emmanuel K. Advantages of early management of facial clefts in Africa. International Journal of Pediatric Otorhinolaryngology 2014;78(3):504–6.

35. Fell MJ, Hoyle T, Abebe ME, et al. The impact of a single surgical intervention for patients with a cleft lip living in rural Ethiopia. Journal of Plastic, Reconstructive and Aesthetic Surgery 2014;67(9):1194–200.

36. Boukhalfa C, Abouchadi S, Cunden N, Witter S. The free delivery and caesarean policy in Morocco: how much do households still pay? Tropical Medicine and International Health 2016;21(2):245–52.

37. Tamanna Keya K, Sripad P, Nwala E, Warren CE. “Poverty is the big thing”: exploring financial, transportation, and opportunity costs associated with fistula management and repair in Nigeria and Uganda. International Journal for Equity in Health 2018;17(70).

38. Gyedu A, Abantanga F, Boakye G, et al. Barriers to essential surgical care experienced by women in the two northernmost regions of Ghana: a cross-sectional survey. 2016;

39. Varela C, Young S, Mkandawire N, Groen RS, Banza L, Viste A. Transportation barriers to access health care for surgical conditions in Malawi; a cross sectional nationwide household survey. BMC Public Health 2019;19(1):264.

40. Kong VY, Aldous C, Clarke DL. Understanding the reasons for delay to definitive surgical care of patients with acute appendicitis in rural South Africa. South African journal of surgery 2014;52(1):2–5.

41. Baker Z, Bellows B, Bach R, Warren C. Barriers to obstetric fistula treatment in low-income countries: a systematic review. Tropical Medicine & International Health 2017;22(8):938– 59.

42. Grimes CE, Henry JA, Maraka J, Mkandawire NC, Cotton M. Cost-effectiveness of surgery in low-and middle-income countries: A systematic review. World Journal of Surgery 2014;38(1):252–63.

43. Chao TE, Sharma K, Mandigo M, et al. Cost-effectiveness of surgery and its policy implications for global health: A systematic review and analysis. The Lancet Global Health 2014;2(6):334–45.

44. Shrime MG, Alkire BC, Grimes C, Chao TE, Poenaru D, Verguet S. Cost-Effectiveness in Global Surgery: Pearls, Pitfalls, and a Checklist. World Journal of Surgery. 2017;41(6):1401–13.

